# Single-tube collection and nucleic acid analysis of clinical samples: a rapid approach for SARS-CoV-2 saliva testing

**DOI:** 10.1101/2021.04.29.21256345

**Authors:** Kyle H. Cole, Alexis Bouin, Caila Ruiz, Bert L. Semler, Matthew A. Inlay, Andrej Lupták

## Abstract

The SARS-CoV-2 pandemic has brought to light the need for expedient diagnostic testing. Cost and availability of large-scale testing capacity has led to a lag in turnaround time and hindered contact tracing efforts, resulting in a further spread of SARS-CoV-2. To increase the speed and frequency of testing, we developed a cost-effective single-tube approach for collection, denaturation, and analysis of clinical samples. The approach utilizes 1 µL microbiological inoculation loops to collect saliva, sodium dodecyl sulfate (SDS) to inactivate and release viral genomic RNA, and a diagnostic reaction mix containing polysorbate 80 (Tween 80). In the same tube, the SDS-denatured clinical samples are introduced to the mixtures containing all components for nucleic acids detection and Tween 80 micelles to absorb the SDS and allow enzymatic reactions to proceed, obviating the need for further handling of the samples. The samples can be collected by the tested individuals, further decreasing the need for trained personnel to administer the test. We validated this single-tube sample-to-assay method with RT-qPCR and RT-LAMP and discovered little-to-no difference between Tween- and SDS-containing reaction mixtures, compared to CDC-approved reagents. This approach significantly reduces the logistical burden of traditional large-scale testing and provides a method of deployable point-of-care diagnostics to increase testing frequency.

## Introduction

The pandemic caused by SARS-CoV-2 (severe acute respiratory syndrome coronavirus 2)^1^ is an ongoing health crisis that has put a spotlight on current methodologies of viral detection. Large scale testing has become a part of the weekly routine to help quell viral spread. In the early stages of the global pandemic, SARS-CoV-2 testing was mainly reserved for those who were symptomatic or had been exposed to other virus-positive individuals, limiting test availability for asymptomatic or pre-symptomatic individuals. Bottlenecks in testing assisted in the spread of SARS-CoV-2 because asymptomatic individuals make up a large portion of the SARS-CoV-2 infected population^2^. While testing has expanded in recent months to include asymptomatic individuals, the length of time between sample collection and result determination has hindered our ability to identify positive individuals before they become infectious and capable of spreading the virus. This lag in test turnaround is due in part to the lengthy sample processing, RNA purification step, and qRT-PCR setup. Furthermore, the high demand for sterile swabs, buffers, tubes, pipette tips and PCR reagents has led to supply chain complications that have further impeded efforts to expand testing capability. Lastly, the number of steps required—from sample collection to result notification—has placed tremendous pressure on clinical labs and staff to avoid mix-ups that could lead to incorrect diagnoses.

Safety is also an important issue, because specimens are potentially infectious during transport until inactivated during processing. While newer methods involving saliva collection have eliminated many of the RNA purification steps, several processing steps remain, and these methods have not had the transformative impact on testing as envisioned. At-home testing solutions, while promising, are still in development and are expensive on a per-test basis. These issues highlight the need for improved testing methods. Current diagnostic-testing workflows for nucleic-acid based methods require long processing times, hindering the time between testing and result determination. Lag in test turnaround is in part due to the RNA purification process necessary to achieve optimal test sensitivity, which has been weighted as the diagnostic standard^3^. While sensitivity of each test is important, testing frequency is also a critical factor for early detection in households and communities, necessitating the development of simple, fast and on-site testing technologies^3,4^. Additionally, the logistics of sample handling and processing requires stringent documentation to maintain sample identity. To address these issues, we have developed a simple, cost-effective, and expedient method of viral testing without pipetting, utilizing reverse transcription and quantitative polymerase chain reaction (RT-qPCR) or reverse transcription and loop-mediated isothermal amplification (RT-LAMP), with the purpose of increasing test simplicity and frequency.

Standard methods of viral diagnostics rely on viral transport media (VTM) to preserve the specimen after collection from the patient. VTM does not inactivate viruses; therefore, patient specimens collected in VTM are potentially infectious. While inactivation is performed by sample heating or by the addition of sodium dodecyl sulfate (SDS)^5^, these steps are performed after transportation and delivery to a diagnostic facility. Recent methods have aimed to streamline this process of viral inactivation and genomic RNA extraction for RT-qPCR^6,7^, RT-LAMP^8,9^, and CRISPR/Cas9^10-12^ by utilizing a heat inactivation step prior to assessment. However, these techniques require further handling of the inactivated samples, such as transfer of specific volumes of the sample material into the diagnostic solution for downstream analysis, thus prolonging the process and requiring dedicated personnel or robotic facilities to handle the samples.

We sought to expedite the inactivation of viruses, bacterium, and human cells by employing the common anionic surfactant, dodecyl sulfate (SDS), for the denaturation of viral and cellular structures and subsequent release of RNA with the addition of ethylenediaminetetraacetic acid (Tris-EDTA, TE) to chelate divalent metal ions required for nuclease activity. Others have utilized similar approaches using surfactants (SDS, polysorbate [Tween], Triton X-100, and nonyl phenoxypolyethoxylethanol [NP-40])^5^, reducing agents (e.g., tris(2-carboxyethyl)phosphine [TCEP], dithiothreitol [DTT]) and EDTA, and/or heat inactivation^8,13^. While the inclusion of SDS at low millimolar concentrations into any reaction mixture will inactivate common pathogenic viruses, it also leads to protein denaturation^14^ and is thus incompatible with enzymatic activity, such as subsequent RT and PCR steps. However, SDS can be sequestered by non-ionic surfactants, such as Tween, at or above their critical micelle concentration (CMC) via hydrophobic interactions^15^. The inclusion of Tween in an enzymatic reaction mixture therefore abrogates the inhibitory effects of SDS, allowing for SDS-treated clinical samples to be added directly to an enzyme reaction mixture.

Current clinical sample procurement methods are accomplished using sterile swabs for the collection of nasopharyngeal or nasal samples, in addition to saliva collection, by utilizing specially fitted collection tubes for direct oral deposition. All these methods require the transportation of the sample for further inactivation and RNA retrieval, prior to performing molecular diagnostics. Both the nasopharyngeal swabs and saliva collection tubes have been plagued by supply chain issues and remain a significant expense. We sought a simple-to-use method of sample acquisition that could be easily mastered by untrained individuals for self-collection. Plastic inoculation loops used in microbiology filled this need, because they are inexpensive, disposable, and can reproducibly remove specific volumes (e.g., 1 µL) of saliva (Figure 1A). These loops provide a means of consistent, small-volume saliva retrieval in addition to their convenient size for introduction into a PCR test tube. Viral samples retrieved by loop can be directly inactivated by applying the sample directly to an SDS/TE mixture and briefly mixing.

**Figure 1:**
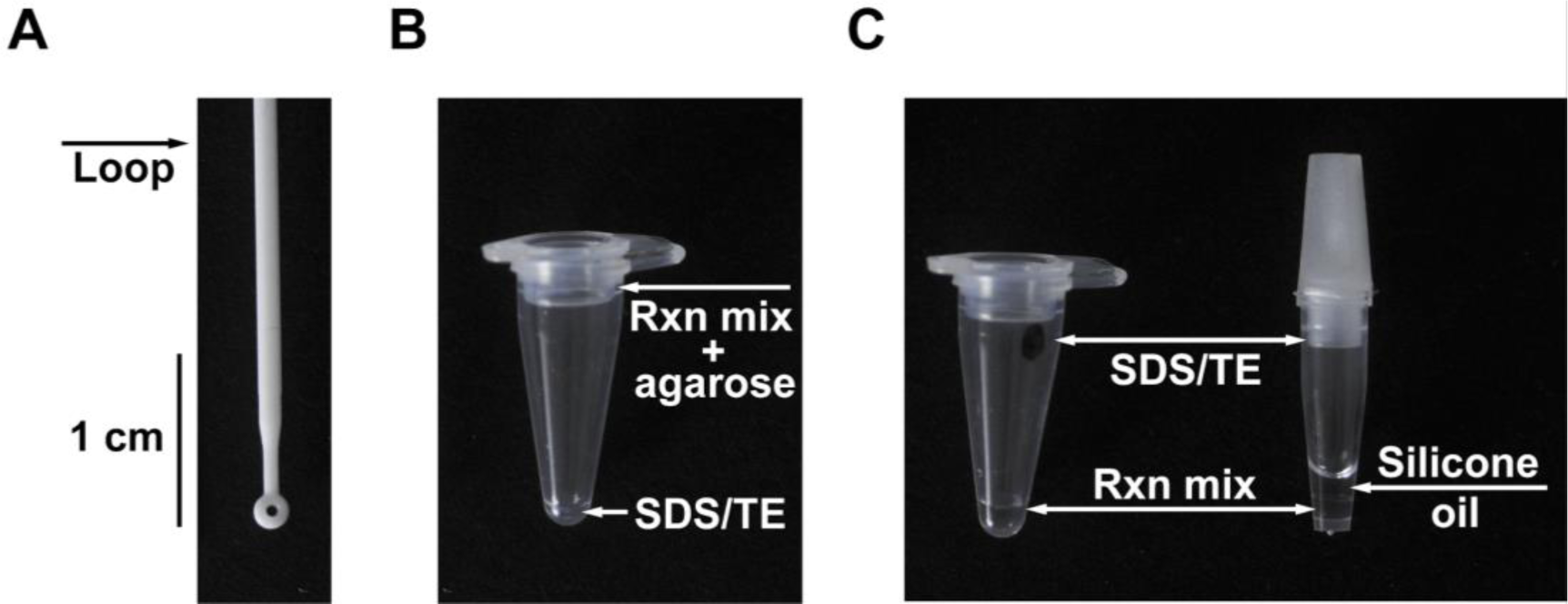
Inoculation loop for sample acquisition and configuration of reaction tubes. (**A**) A 1 µL inoculation loop is used for oral sample withdrawal and collection. The loop fits within 0.1 mL optically transparent PCR tubes. **B**) A 0.1 mL PCR tube with a Tween-containing reaction mixture immobilized by agarose is placed inside the cap of the tube and 1 µL of SDS/TE solution is placed at the bottom. A clinical sample withdrawn using the inoculation loop is transferred to the bottom of the tube and denatured by mixing it with the SDS/TE solution using the same loop. Subsequently, the enzyme reaction mixture containing all components necessary for nucleic acid detection and Tween 80 is transferred from the cap to the bottom of the tube by centrifugation or shaking. (**C**) Alternatively, 0.1 mL PCR tube with a dried 1 µL spot of SDS/TE on the sidewall of the tube and 5 µL of a Tween-containing reaction mixture at the bottom can be used (left tube). The inoculation loop is rubbed on the side of the tube to dissolve the SDS/TE mixture and denature the clinical sample. The same loop is then used to drive the sample to the bottom of the tube, where it mixes with the nucleic acid detection mixture. For systems with optical readout at the bottom of the sample tube (e.g., Biomolecular Systems Mic, Quantabio Q tube or Qiagen RotorGene Q) the SDS/TE solution can be dried on the sidewall and the Tween-containing RT-qPCR reaction mixture can be stored underneath 5 µL of silicone (or mineral) oil (right tube).

To utilize this system within a single tube, the SDS/TE solution must be initially separated from the enzyme reaction mixture. This was accomplished by immobilizing the reaction mixture with agarose in the lid of the tube, while supplying the SDS/TE mixture at the bottom (Figure 1B). Alternatively, the SDS/TE could be dried on the inner-side wall of the PCR tube with the reaction mix contained at the bottom of the PCR tube (Figure 1C). This configuration allows for streamlined sample retrieval, inactivation by SDS/TE, and subsequent introduction into the Tween-containing enzymatic reaction mixture. Utilizing this approach, we demonstrate both RT-qPCR and RT-LAMP analyses of viral particles harboring the SARS-CoV-2 spike protein. The method thus represents a practical sample-to-assay approach with the potential to accelerate the diagnostic process as well as providing a cost-effective solution for increasing testing frequency.

## Results

### Optimization of SDS/Tween solutions

We first tested qPCR reactions to determine the concentration of Tween necessary to sequester SDS and prevent the inhibition of amplification. To test this, human RNase P gene (RPP30) was amplified from a control plasmid at varying concentrations of Tween 20 and Tween 80. We first established that 0.005% (w/v) final concentration of SDS is sufficient to inhibit the amplification reaction. We then increased the SDS concentration to 0.01 % (w/v) and tested Tween 20 and Tween 80 at 1%, 2%, and 5% (v/v) for the ability to amplify DNA (Figure 2A). At 5% (v/v) Tween 20 supported amplification in the presence of SDS but was ineffective at 1% and 2% (v/v). Tween 80 abolished the inhibitory effects of SDS at all three concentrations and supported amplification comparable to the control reaction. The results are consistent with the higher stability of the Tween 80 micelles, compared to Tween 20 (Tween 20 has a higher critical micelle concentration than Tween 80)^16^. We chose to use 3% (v/v) Tween 80 for our subsequent applications.

**Figure 2:**
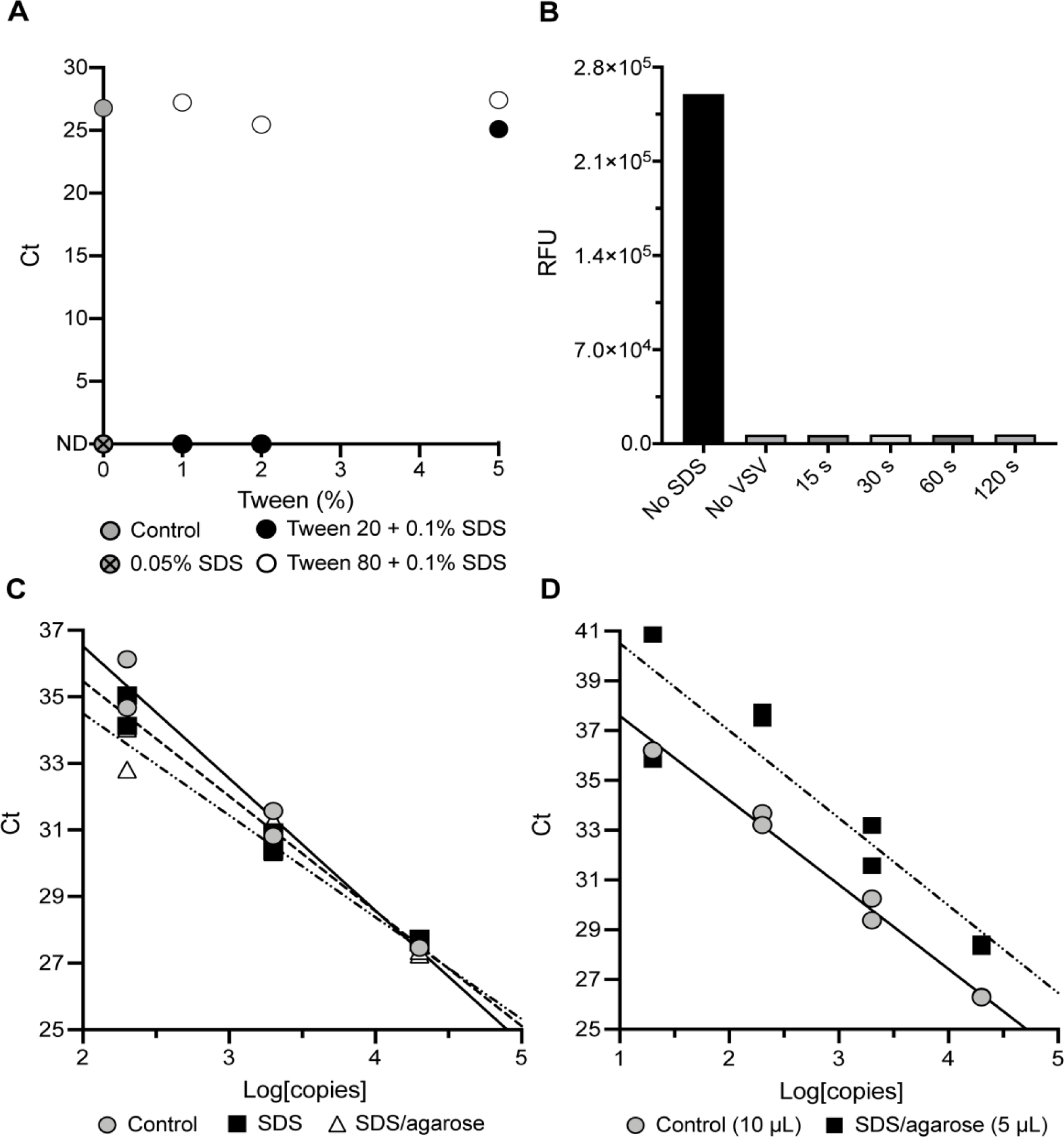
Denaturation of live viruses and nucleic acid amplification reactions using soap/surfactant combinations. (**A**) Determination of the minimal Tween 20 and Tween 80 concentration necessary for SDS sequestration to enable amplification of 20,000 copies/µL of RNase P DNA. ND: not detected. (**B**) Fluorescence assay of eGFP-expressing recombinant VSV-SARS-CoV-2 (10^7^ pfu/mL) infecting Vero-E6 cells after incubation with 0.1% (w/v) SDS for 15–120 seconds. Only a sample that was not exposed to SDS showed infectivity. 15 sec exposure to 0.1 % SDS proved sufficient to completely inactivate the virus, likely due to dissolution of its membrane and denaturation of the sample proteins. (**C**) Comparison of 10 µL qPCR reactions for SARS-CoV-2 *N1* gene DNA amplification, containing 3% (v/v) of Tween 80 with and without 0.01% SDS (w/v, final) and 1% (w/v) agarose. (**D**) Comparison of a 10 µL qPCR reaction for SARS-CoV-2 *N1* gene DNA containing 3% (v/v) Tween 80 with a 5 µL qPCR reaction for SARS-CoV-2 N1 DNA detection, containing 3% (v/v) Tween 80, 0.01% SDS (w/v, final) and 1% (w/v) agarose. In all conditions containing SDS, the DNA samples were initially incubated with 0.1% SDS (w/v) in Tris/EDTA buffer to simulate denaturing conditions for clinical samples. The samples were then centrifuged to mix with the Tween-containing PCR reactions.

A vesicular stomatitis virus (VSV) expressing enhanced green fluorescent protein (eGFP) and the SARS-CoV-2 spike protein (VSV-eGFP-SARS-CoV-2^17^), was used as a model system to test the SDS inactivation efficacy. Like SARS-CoV-2, VSV is an enveloped RNA virus that loses infectivity when the envelope is disrupted by detergent treatment. To assess virus inhibition, 1-µL inoculation loops were dipped into a VSV-eGFP-SARS-CoV-2 viral suspension of 10^7^ pfu/mL and introduced to a dried 1 µL spot containing 0.1% (w/v) SDS in a Tris-EDTA buffer on the inner-side wall of a PCR tube. The 1 µL viral samples on the inoculation loops were mixed on the SDS spot for 15-120 seconds prior to introduction into a 5-µL RT-qPCR reaction mixture containing 3% (v/v) Tween 80 at the bottom of the PCR tube. To prevent cross contamination, a new 1-µL loop was used to remove 1 µL from the RT-qPCR reaction mixtures and inoculate Vero-E6 cell monolayers. Cells were seeded on 6-well culture plates 24 hours prior to infection. Cells were grown for 5 days at 34 °C under 5% CO2 with daily fluorescent signal monitoring and until the untreated viral sample achieved complete infection. After incubation, eGFP expression in SDS-treated, no-SDS, and no-VSV samples was measured on a CLARIOstar Plus Microplate Reader (Figure 2B). Inhibition of VSV-eGFP-SARS-CoV-2 was observed at the minimum time of mixing on the 0.1% (w/v) SDS/TE spot at 15 seconds and was comparable to mixing for longer periods and to the no-VSV control. Far greater eGFP expression was observed in the untreated VSV-eGFP-SARS-CoV-2 control. The significant fluorescence observed in the untreated sample demonstrates that SDS fully inactivates the virus, but the 3% (v/v) Tween 80 dissolved in the enzyme reaction mixture does not. These results illustrate that 0.1% (w/v) SDS with 15 seconds of mixing is sufficient to inactivate VSV-eGFP-SARS-CoV-2 and further shows that additional heat-inactivation is unnecessary for virus inactivation.

For point-of-care testing, immobilization of the master mix is necessary to prevent the mixing of the Tween-80-containing enzyme/primer master reaction with the SDS, particularly when SDS is supplied at the bottom of the reaction tube. Several methods can be used for the separation of the reagents within a single tube: 1) mineral oil or silicone oil can be overlaid above the enzyme mix^18^ (Figure 1C), 2) low-melting temperature wax can be overlaid on top of the mixture and solidified to make a solid barrier at room temperature^19^, or 3) low-melting-point agarose can be introduced into the mixture at a slightly elevated temperature (e.g., 42 °C) and then stored at room temperature or lower to immobilize the mixture. We confirmed that both the oil overlay and the agarose inclusion supported immobilization of the enzyme mixture, and for the majority of experiments, we used low-melting-point agarose for its ability to melt at the low reaction temperatures (≤50 °C), enzyme stabilization^20^, and its optical transparency^21^. The addition of 1% (w/v) agarose in the presence of SDS (0.01% (w/v) final) and 3% (v/v) Tween 80 showed no discernable inhibition of the amplification of SARS-CoV-2 *N* gene from a SARS-CoV-2 plasmid when compared to reactions containing 3% (v/v) Tween 80, with and without 0.01% SDS (w/v, final) in 10 µL samples (Figure 2C).

Next, we compared a 5 µL reaction containing the SDS/Tween/agarose to a standard 10-µL reaction mixture containing 3% (v/v) Tween 80 to test any inhibitory effects under more stringent conditions (Figure 2D). The smaller reaction volume with the addition of both 0.01% SDS (w/v, final) and 1% (w/v) agarose increased the number of cycles necessary for detection of SARS-CoV-2 N1 at each concentration of plasmid DNA, thus exhibiting a slight decrease in amplification efficiency (Figure 2D). Importantly, a 5 µL reaction with a sample introduced into a 1 µL drop of SDS solution and subsequently mixed with a master mix containing Tween 80 allowed quantitative detection of DNA plasmids, providing a facile and economical approach to the analysis of clinical samples in a single tube.

### RT-qPCR of SARS-CoV-2 viral particles and synthetic RNA

While the SDS/Tween assay was compatible with qPCR, understanding RT-qPCR functionality was necessary to establish a method for sample-to-assay assessment of infection. To assess RT-qPCR, 50 µL of two positive SARS-CoV-2 patient nasal swabs samples stored in VTM with 0.5% (w/v) SDS were purified using a Zymo Research Quick DNA/RNA-viral extraction kit and eluted in an equivalent volume of nuclease-free water. We diluted the unpurified RNA (VTM with 0.5% [w/v] SDS) five-fold in nuclease-free water to reach an effective SDS concentration of 0.1% (w/v) and compared that to equally dilute purified samples (Figure 3A). Reactions were performed at 10 µL with the RT-qPCR reaction mixture immobilized in the lid of the reaction tube (0.1 mL optically transparent qPCR tubes). The enzyme mixture also contained SARS-CoV-2 N1 primers and probe, 3% (v/v) Tween 80, and 0.5% agarose (w/v). Samples were prepared in triplicate with 1-µL of sample added by pipetting. The samples were then placed in a 96-well tube frame and centrifuged for 30 seconds on a benchtop centrifuge at 3000 × *g*. The frame was placed into a Bio-Rad CFX Connect thermal cycler and the RNA was detected using the CDC-recommended amplification protocol: 25 °C for 2 minutes, 50 °C for 15 minutes, 95 °C for 2 minutes, and 45 cycles of 95 °C for 3 seconds, and 55 °C for 30 seconds. Amplification of both purified and unpurified patient samples provided consistent Ct values with no observable differences between them.

**Figure 3:**
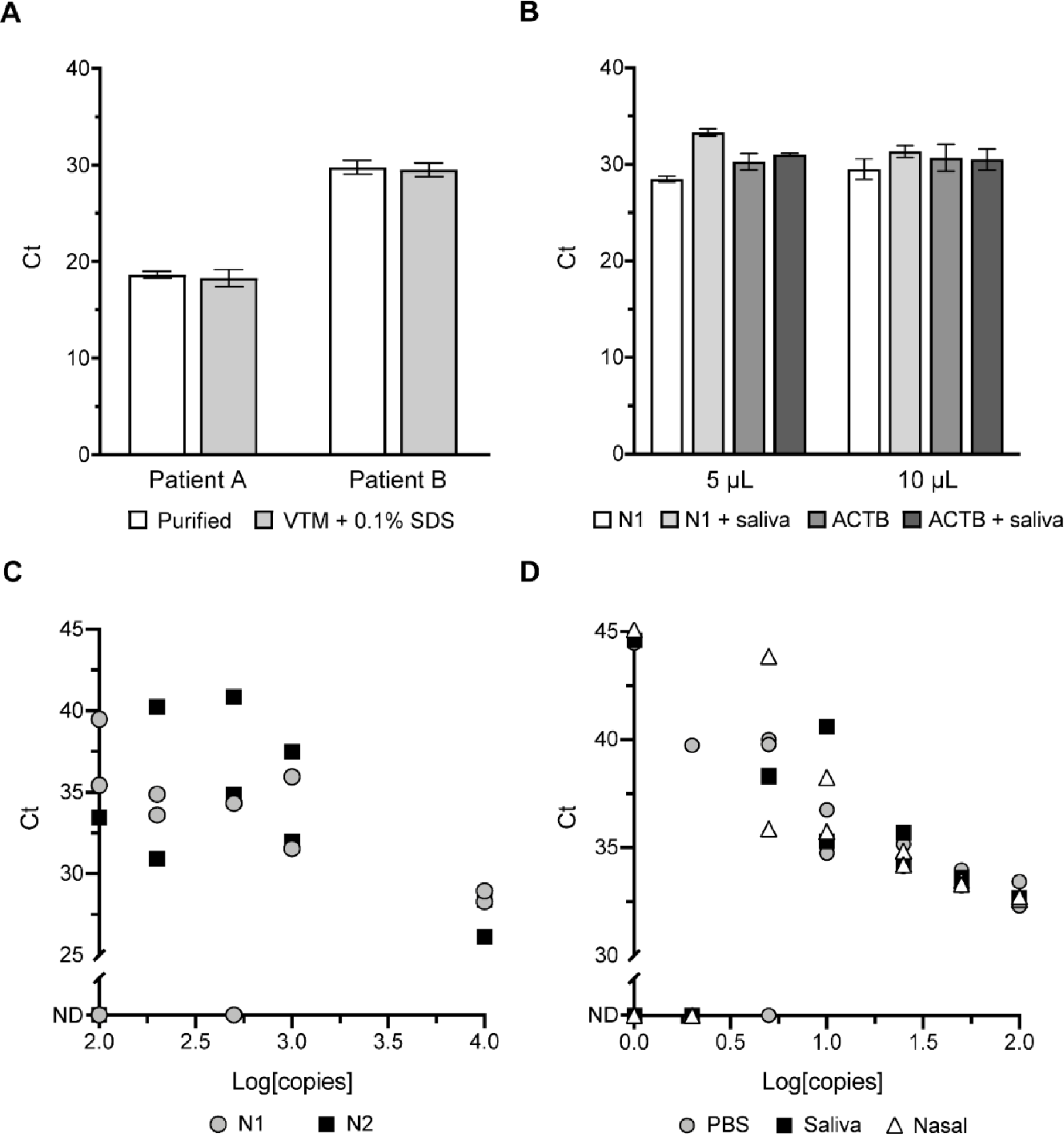
Single-tube sample-to-assay RT-qPCR of SARS-CoV-2 positive-patient RNA sample and inactivated viral particles. (**A**) Comparison of five-fold diluted purified and unpurified SARS-CoV-2 patient RNA samples (nasal swabs stored in VTM + 0.5% [w/v] SDS) in triplicate 10 µL RT-qPCR reactions containing SARS-CoV-2 N1 primers and probe, 3% (v/v) Tween 80, and 0.5% (w/v) agarose. Samples were diluted five-fold to achieve a 0.1% (w/v) SDS working concentration for unpurified samples in VTM. (**B)** Comparison of 5 µL and 10 µL RT-qPCR reactions containing 3% (v/v) Tween 80, and 0.5% (w/v) agarose with a purified SARS-CoV-2 patient RNA sample. Triplicate reactions were performed at each volume, comparing the patient sample with and without spiking into saliva. Reactions were performed with SARS-CoV-2 N1 and beta-actin (ACTB) primer-probe sets. (**C**) RT-qPCR results of SARS-CoV-2 synthetic RNA (n = 11) in water. RT-qPCR was performed for 45 cycles using SARS-CoV-2 N1- and N2-specific primers and probes on a Quantabio Q thermal cycler. Samples were tested blind with concentrations and storage media revealed thereafter. The *y*-axes are truncated to indicate results with no detectable amplification (ND). (**D**) Scatter plot of Ct value (*y*-axis) versus log[copies] of SARS-CoV-2 viral particles (*x*-axis) of inactivated SARS-CoV-2 viral particle samples (n = 49) in PBS, saliva, and nasal media. RT-qPCR was performed for 45 cycles with SARS-CoV-2 N1-specific primers and fluorescent probe on a Bio-Rad CFX Connect thermal cycler.

We then assessed one purified patient RNA sample that was spiked into saliva obtained from one of our authors to determine whether saliva could inhibit or interfere with detection (Figure 3B). Reactions were performed at two volumes (5 µL and 10 µL) and contained SARS-CoV-2 N1 and beta-actin (ACTB) primer-probe sets, 3% (v/v) Tween 80, and 0.5% agarose (w/v). Samples were prepared in triplicate, retrieved with 1-µL inoculation loops, and inactivated by mixing with 0.1% (w/v) SDS in TE and 5 mM DTT^8^ stored at the bottom of the reaction tube. Reactions were performed using the previously mentioned CDC-recommended protocol. A small difference in amplification of SARS-CoV-2 N1 was observed at both reaction volumes when the RNA sample was spiked into saliva. However, this same difference was not observed for ACTB between either the 5 µL or 10 µL reaction volumes. We hypothesize that this difference is due to the purified-patient RNA degrading when incubated in saliva, prior to inactivation by SDS. The ACTB mRNA was likely protected within cells collected with saliva and released after SDS treatment, explaining the consistency of the Ct values in the ACTB reactions with and without saliva. Overall, these results showed that the reaction mixture containing both Tween 80 and agarose can detect SDS treated SARS-CoV-2 RNA.

Next, we tested our sample-to-assay method utilizing an RT-qPCR reaction mixture overlaid with silicone oil. We tested samples of synthetic SARS-CoV-2 RNA stored in water (n=11, provided by the XPRIZE Foundation; Figure 3C). The RT-qPCR reaction mixture was prepared and stored beneath a layer of silicone oil in a Quantabio Q tube produced specifically for the Q qPCR instrument, which measures fluorescence from the bottom of the tubes by rotating the samples above immobile excitation LEDs and photomultiplier detectors. This process also serves to centrifuge the samples to the bottom of the tubes. We assessed these samples by first collecting each sample using an inoculation loop and mixing the sample on a dried SDS/TE spot on the side of the tube for 15 seconds. After mixing, the sample was driven by the inoculation loop to the bottom of the test tube (past the silicone oil) into the reaction mixture. The samples were incubated at 37 °C for 30 seconds, 42 °C for 4 minutes, 50 °C for 5 minutes to promote cDNA synthesis and subsequently PCR-amplified using the QuantaBio Q qPCR recommended conditions: 95 °C for 1 minute, and 45 cycles of 95 °C for 5 seconds, and 55 °C for 10 seconds. Because the samples consisted of synthetic RNA stored in water, it was expected that some of the synthetic SARS-CoV-2 RNA had degraded. This experiment provides an example of this method when utilizing a system that reads fluorescence from the bottom of the tube and maintains partitioning of the reaction mixture with silicone oil. Additionally, this provides a useful method of detection of >200 copies/µL of viral RNA.

We next assessed 49 inactivated SARS-CoV-2 viral particle samples diluted in phosphate buffered saline (PBS), saliva, and nasal media, provided as blind samples by the XPRIZE Foundation (Figure 3D). Reaction tubes (0.1 mL optically transparent qPCR tubes) were prepared by drying 1 µL of 0.1% (w/v) SDS in TE buffer solution on the inner-side wall of the PCR tube. Samples were collected by dipping 1-µL inoculation loops in the stored samples, swirled on the dried SDS for 15 seconds and subsequently driven to the bottom of the tube containing a 5-µL RT-qPCR reaction mixture that included 3% (v/v) Tween 80 and N1-specific primers and probe. The sample tubes were placed in a 96-tube frame and centrifuged for 30 seconds on a benchtop centrifuge at 3000 × *g*. The frame with the sample tubes was then placed in a Bio-Rad CFX thermal cycler and the RNA was detected using the CDC-recommended amplification protocol: 25 °C for 2 minutes, 50 °C for 15 minutes, 95 °C for 2 minutes, and 45 cycles of 95 °C for 3 seconds, and 55 °C for 30 seconds. Reactions containing 100, 50, and 25 copies of SARS-CoV-2 viral particles appeared at similar cycles regardless of storage media (n = 18) while samples containing 10 copies varied between 35 and 40 cycles (n = 7) with greater variability and false-negative results at lower concentrations (Figure 3D). Based on these results, we estimate our limit-of-detection (LOD) to be 10 viral genome copies/µL.

### Sample-to-assay RT-LAMP with the addition of Tween 80 and SDS

After verifying sensitive, reproducible RT-qPCR detection with SDS-denatured samples and stabilization of the enzymatic reactions with Tween, we then tested RT-LAMP for a single-tube sample collection and analysis. RT-LAMP has key advantages over RT-qPCR because it is performed isothermally and does not require additional equipment for readout, resulting in diagnostic simplicity. These advantages make RT-LAMP a promising tool for point-of-care detection of viral infection^8,9,22-26^. Previous work has established that RT-LAMP can withstand 3% (v/v) Tween 20 in addition to 3% (v/v) Triton X-100 with little inhibition of amplification^8^. We determined that 1-3% (v/v) Tween 80 was sufficient to sequester 0.1% (w/v) SDS for amplification by RT-LAMP (data not shown). For convenience, we utilized the New England Biolabs (NEB) Isothermal Amplification Buffer containing 0.1% (v/v) Tween 20 and supplemented it with 0.9% (v/v) Tween 80 for our assays.

To validate RT-LAMP detection of RNA samples in the presence of SDS and Tween, real-time fluorescence measurements were collected on a real-time PCR thermal cycler, with the addition of the nucleic-acid intercalating dye SYTO 82^27-29^. Four concentrations of SARS-CoV-2 viral particles (100, 50, 25, and 10 copies/µL) were collected using 1 µL inoculation loops and incubated at 65 °C for 50 minutes in the RT-LAMP reaction. Amplification of 100 to 10 copies of RNA was detected after 33 minutes using N1-specific primers designed by Huang *et al*. (Figure 4A)^30^. Non-specific amplification was not observed in no-reverse-transcriptase (NRT) and no-template (NTC) controls, which was further confirmed by agarose gel electrophoresis (Figure 4B).

**Figure 4:**
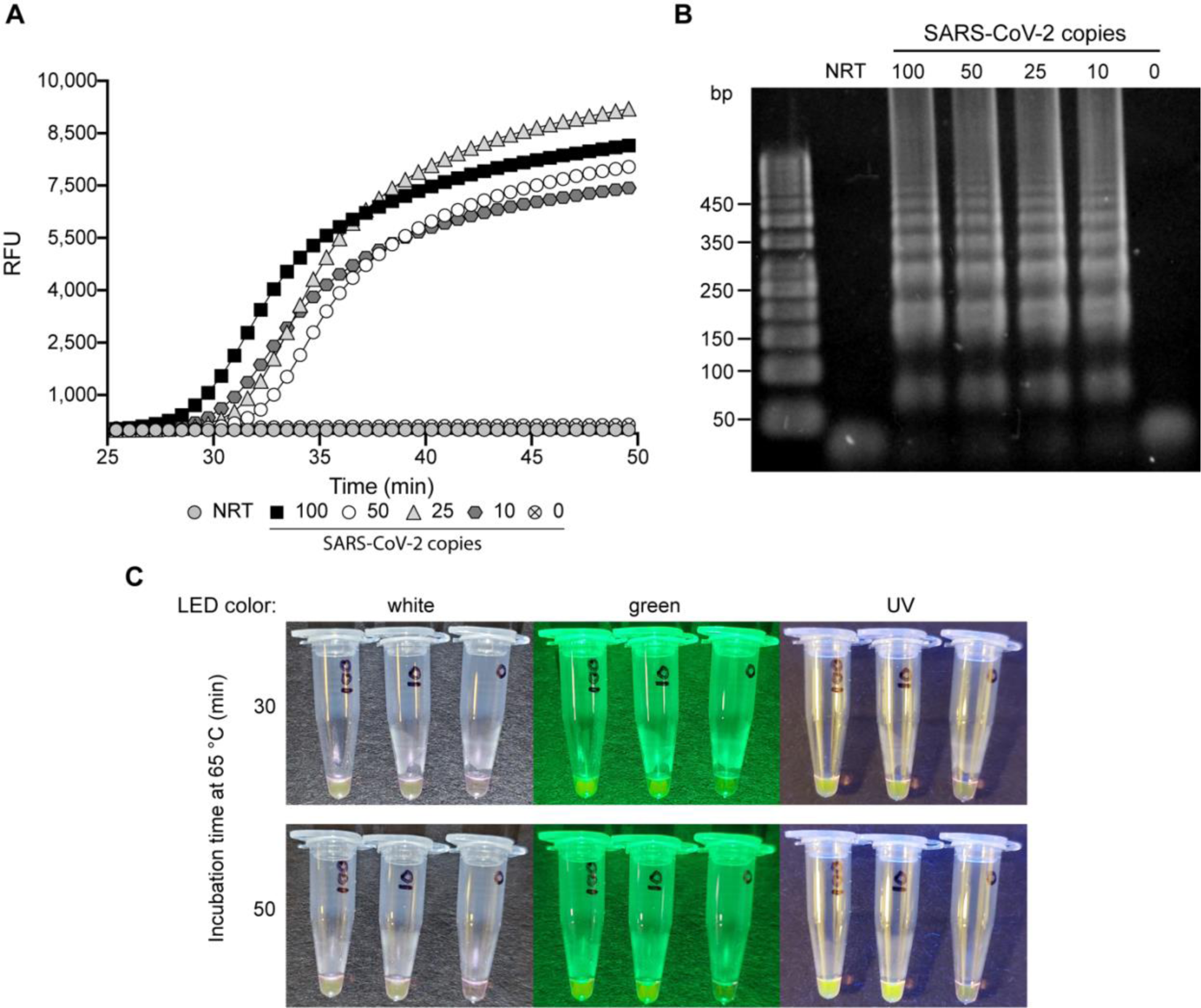
RT-LAMP amplification with SDS and Tween 80. (A) Real-time fluorescence of SARS-CoV-2 *N* gene amplification by RT-LAMP was performed at 65 °C for 50 minutes. Indicated concentrations (copies) of SARS-CoV-2 viral particles were assessed including a no-reverse-transcriptase (NRT) and no-template control (NTC; 0 copies), with all SARS-CoV-2 samples providing a positive result after ∼30 minutes. (B) Amplification products (1 µL) from the real-time fluorescence RT-LAMP reaction were fractionated using agarose gel electrophoresis. The expected laddering pattern of LAMP amplification products was observed at all SARS-CoV-2 viral particle concentrations and not observed in NRT and NTC controls. (C) Fluorescence of RT-LAMP reactions of SARS-CoV-2 viral particles containing 20 µM SYTO 82 observed after 30 and 50 minutes of incubation at 65 °C. Reactions were analyzed with white, green, and UV (365 nm) handheld LED flashlights at each time point, with readily observable fluorescence of amplification reactions starting from 100 copies and 10 copies (labeled) after 30 minutes with each illumination. After 50 minutes, fluorescence in the 100-copy and 10-copy tubes increased with no observable amplification in the 0-copy control.

As mentioned, the isothermal reaction of RT-LAMP eliminates the need for expensive detectors, and with additional indicators, positive results can be observed without costly equipment or gel electrophoresis. Previous methods have utilized pH (e.g., phenol red) or complexometric dyes (e.g., hydroxynaphthol blue), and nucleic acid intercalators (e.g., SYBR Green, EvaGreen, and SYTO 82) for the detection of target RNA^8,9,22,23,28,31,32^. After validating RT-LAMP using real-time fluorescence, we hypothesized that amplification can be observed without dedicated fluorescence readers. We tested this by increasing the concentration of SYTO 82 in the RT-LAMP reaction from 5 to 20 µM. Amplification of 100 copies and 10 copies of SARS-CoV-2 RNA was readily observed in 30 minutes at 65 °C, with no amplification observed in the NTC reaction after 50 minutes (Figure 4C). These results were obtained under consumer-grade white, green, and UV (365 nm) excitation light using handheld LEDs and imaged by a mobile phone.

## Discussion

We present a method aimed at increasing the frequency of viral testing by decreasing the handling and time between sample collection and assessment. This methodology provides a simple approach to virus inactivation for direct RNA detection in patient saliva within a single reaction tube (Figure 1). In previous studies, nucleic acid detection techniques have required extraction of the genetic material from the clinical samples^8,10,11,30,33,34^, with some methods using heat denaturation to inactivate the pathogens and human cells in the clinical samples and release the genetic material for subsequent analysis^6-9,12,26,35,36^. In most cases, the sample collection and inactivation or extraction is performed in separate tubes from the analysis step and thus require additional handling by trained personnel. We sought to simplify the sample preparation to decrease the time and effort necessary to collect, denature, and analyze the sample. In effect, we have eliminated all pipetting and open-tube processing steps. If an individual collects their own specimen, adds it into the tube, and caps it, trained staff will only need to place the tube in the RT-qPCR instrument and analyze the results. After sample collection, the tube can be washed on the outside with a denaturing solution (e.g., soap or sanitizer) and placed in a tube rack or directly into the qPCR instrument by the tested individual, further eliminating handling by technical personnel.

We first standardized the collection of the clinical samples, so that the samples can be directly analyzed in small volumes used in biochemical analyses, such as RT-qPCR, RT-LAMP, or CRISPR-type experiments. We further simplified the handling and safety of the collected samples and to avoid unnecessary steps that require exposure of technicians to potentially infectious samples and add expense and time to the analysis.

In laboratory settings, different types of disposable and mechanical pipettes are used for measuring precise volumes of liquid, particularly in the microliter to milliliter range. To our knowledge, disposable pipettes capable of dispensing microliters of clinical samples do not exist and mechanical pipettors are expensive and require training for correct use. Therefore, we adopted standard 1 µL inoculation loops (Figure 1) commonly used in microbiological experiments to collect a relatively precise volume of saliva. In our experience, non-experts easily mastered saliva collection from their oral cavity (e.g., as a cheek swab) using these loops. Saliva is more viscous than water, making the sample collection and transfer easier than standard buffers or culture media.

To render the clinical samples noninfectious, we used laboratory detergent (SDS), which is commonly used to denature proteins and disintegrate lipid membranes. Detergent is the most active and widely available denaturant and, as expected, we observed that incubating a recombinant VSV-eGFP-SARS-CoV-2 virus sample for 15 seconds made the sample completely non-infectious (Figure 2B). However, SDS also denatures enzymes that are used for nucleic acid detection, and to avoid a purification step, we required a mechanism of SDS sequestration from the solution. One approach is to precipitate the dodecyl sulfate with potassium, which, unlike the sodium salt, is insoluble; however, precipitation would lead to flocculates that could potentially hamper the downstream analysis of the solution. Instead, we chose to adsorb dodecyl sulfate into micelles formed by very stable surfactants, such as the Triton and Tween series of non-ionic detergents. Of the commercially available non-ionic detergents, Tween 80 forms the most stable micelles with low micromolar critical micelle concentration over a wide range of temperatures. Both Tween and Triton surfactants have been widely used in enzymatic reactions, including PCR and droplet PCR, and are therefore highly suitable for SDS sequestration. With the addition of Tween to an RT-qPCR reaction, the denaturing effects of the SDS are abolished, enabling amplification of target DNA (Figure 2A). This protocol provides an expedient approach to viral RNA liberation within a PCR tube containing the reaction mixture and decreases the time and effort required for diagnosis.

The addition of agarose was tested to determine whether a stabilizing medium could be supplemented to better immobilize the reaction mixture away from the place where a small volume of SDS/TE was applied (and sometimes dried). We propose this approach as a means of immobilizing the reaction mixture during sample collection to prevent the loss of the reagents crucial for diagnostics. We observed no inhibition of qPCR due to SDS/Tween and agarose at equal reaction volumes (10 µL), when compared with standard reaction conditions (Figure 2D), and the amplification efficiency was only slightly lower when 1 µL samples were incubated in 5 µL enzyme reactions. A similar result was observed for RT-qPCR utilizing a column-purified SARS-CoV-2 patient sample, even in the presence of saliva (Figure 3B). We believe that this slight decrease in amplification efficiency is acceptable because the mean cycle threshold of N1 primer sets used in standard RT-qPCR protocols for self-saliva collection is 32.8 for asymptomatic patients and those with early- and late-onset of disease^37^.

Testing inactivated SARS-CoV-2 viral particles in PBS, saliva, and nasal media revealed variation at concentrations of 10 copies and below, giving us an estimated limit of detection of 10 copies/µL (Figure 3D). Because others have illustrated a detection limit of 5.6 copies/µL of SARS-CoV-2 following the Centers for Disease Control and Prevention’s RT-qPCR guidelines for the N1-primer set, we believe our protocol falls within a meaningful detection range for this coronavirus^38^. This slight sensitivity decrease is compensated by the great reduction in diagnostic handling and turnaround time, thereby allowing for more frequent testing. Furthermore, 10 copies/µL is almost two orders of magnitude below the median viral load of sputum samples (752 copies/µL) collected from infected individuals^39^.

RT-LAMP has the potential to fill a niche previously occupied by rapid-antigen tests. It has been reported that rapid-antigen tests have a high false-negative rate in saliva samples with Ct ≥25 while RT-LAMP had greater sensitivity for samples with Ct ≤35^37^. Our method applied to RT-LAMP allowed for the detection of 10 copies (mean Ct = 34.8) of SARS-CoV-2 N1 RNA in 30 minutes (Figure 4). By increasing the concentration of SYTO 82 in the RT-LAMP reaction, a positive result could be seen under white light and better observed with green and UV (365 nm) LEDs. Based on these results, RT-LAMP would require the same incubation time as commercial rapid-antigen tests (30 minutes) with greater sensitivity, although requiring incubation at 65 °C. Additionally, our method of RT-LAMP does not require the addition of buffering reagents typically required of lateral-flow rapid-antigen tests. While it does not have the same accuracy of RT-qPCR and suffers from non-specific amplification activity, RT-LAMP paired with this sample-to-assay method could replace the market of rapid-antigen tests while maintaining the simplicity of result determination.

Our testing strategy has several cost-saving measures. Inoculation loops are inexpensive, with retail prices at about 2¢ (0.02 USD). The use of 5-µL enzyme reaction volumes is 4 times less expensive than the 20-µL reactions recommended by the CDC, and thus the cost of qRT-PCR master mix and primers is 4 times less. All pipetting steps have been removed, eliminating the need for pipettes and disposable tips, and only a single PCR tube is needed per test. SDS, Tween-80, and low melt agarose are all inexpensive at the quantities needed. Technician time and skill is reduced to transferring tubes to the PCR instrument, and even this step can in principle be performed by the tested individuals; therefore, minimal specialization is required to setup and administer a testing unit based on our method. In summary, we estimate a cost of ∼0.5 USD per test, not counting fixed costs, such as the cost of the qPCR instrument.

## Conclusion

Our single-tube clinical sample analysis method greatly improves sample collection and detection in situations that require frequent and widespread testing, such as during a pandemic. When large-scale testing is required, as exemplified by the SARS-CoV-2 global pandemic, bottlenecks in sample collection, extraction, and test run-times are exacerbated. By combining viral inactivation with RNA release and detection in a single reaction tube, turnaround of test results can be significantly shortened. Samples are extracted and added directly to the reaction mixture by patients, eliminating the need for intermediary interaction by medical workers at testing sites. Further shortening of turnaround time results from the decreased need for specialized equipment and trained personnel when analyzing the collected samples, allowing for testing that requires only PCR instruments, some of which are stable enough to work dependably in mobile testing units, or, as in the case of RT-LAMP detection, just LED flashlights. The inclusion of unique QR or barcodes to individual test tubes would allow anonymization of the samples and when coupled with smart-phone registration, an expedient method of reporting of results. By reducing the intermediary steps required for nucleic acid extraction, mobile testing facilities will have the ability to test samples on-site. We envision the deployment of mobile diagnostic facilities used at populous institutions such as schools, hospitals, churches, transportation hubs, and manufacturing facilities, and thereby providing near-real-time information about infection rates.

## Data Availability

Tabulated data is available for use through the corresponding author.

## Acknowledgements

We thank Paul W. Rothlauf and Sean P.J. Whelan for the recombinant VSV-eGFP-SARS-CoV-2 and the XPRIZE Foundation for providing inactivated SARS-CoV-2 viral particles through the XPRIZE Rapid Covid Testing competition. The authors wish to acknowledge the support of the Chao Family Comprehensive Cancer Center Experimental Tissue Shared Resource, supported by the National Cancer Institute of the National Institutes of Health under award number P30CA062203. The content is solely the responsibility of the authors and does not necessarily represent the official views of the National Institutes of Health. The project described was supported by the UC Irvine COVID-19 Basic, Translational and Clinical Research Funding Opportunity (to A.L. and B.L.S.), NSF CBET 1804220 (to A.L.), NIH grant AI145003 (to B.L.S.), and by numerous donors through a crowdsourcing campaign (ZotFunder “Simple and Rapid COVID-19 Diagnostic Test”), whose support is gratefully acknowledged. A.L. is a Fellow of the J. S. Guggenheim Memorial Foundation.

## Methods

### Cell and virus culture

Vero-E6 cells (ATCC, CRL-1586) were grown in Dulbecco’s Modified Eagle Medium (DMEM; Gibco, 12800017) supplemented with 10% fetal bovine serum (FBS; Omega Scientific, FB-12) and a 1X antibiotic-antimycotic solution (Omega Scientific, AA-40) and maintained at 37°C with 5% CO2. Vero-E6 cells were seeded 24 hours prior to infection in 6-well plates. Infection was performed using VSV-eGFP-SARS-CoV-2 (generously provided by Paul W. Rothlauf and Sean P.J. Whelan at Washington University in St. Louis^15^) expressing SARS-CoV-2 spike protein (*S* gene). Following infection, cells were grown at 34 °C with 5% CO2 in DMEM supplemented with 10% FBS.

### Fluorescent VSV inactivation assay

Sodium dodecyl sulfate (SDS; Sigma Aldrich, 436143) inactivation was performed by dipping a 1 µL inoculation loop (Globe Scientific, 2810) in a VSV-eGFP-SARS-CoV-2 suspension at 10^7^ pfu/mL and applying the inoculation loop to a dried 1 µL spot of SDS/TE (0.1% w/v SDS, Tris-HCl, pH 8.0) on the side wall of a PCR reaction tube, followed by mixing for 15-120 seconds. The treated inoculation loop was then driven down into a RT-qPCR reaction mixture (see standard RT-qPCR methods) containing 3% (v/v) polysorbate 80 (Tween 80; Sigma Aldrich, P1754) and disposed of. A new 1 µL inoculation loop was then used to sample the inoculated RT-qPCR reaction mixture and introduced into the culture media of Vero-E6 cells seeded in a 6-well culture plate. Cells were grown at 34 °C with 5% CO2 in DMEM supplemented with 10% FBS for 5 days. Cells were fixed in PBS supplemented by 3.7% formaldehyde for 20 minutes at room temperature and washed 3 times with phosphate buffered saline (PBS). Fluorescence was measured using a CLARIOstar Plus Microplate Reader (BMG Labtech).

### DNA and RNA controls

Human RNase P (RPP30) was amplified from the Hs_RPP30_Positive control plasmid (Integrated DNA Technologies, IDT, 10006626). SARS-CoV-2 N1 DNA was amplified from the 2019-nCoV_N_Positive control plasmid (IDT, 10006625). SARS-CoV-2 viral particles and synthetic RNA were provided by the XPRIZE Foundation (Team ID: 3650) and produced by ZeptoMetrix and Twist Biosciences, respectively. Viral particles were stored at 4 °C per manufacturer recommendation. Synthetic RNA was stored at -80 °C per manufacturer recommendation.

### RT-qPCR primer design

Human RNase P (RPP30) primers and probe were synthesized by IDT (oligonucleotides: 25 nmol, standard desalting; hydrolysis probe: 25 nmol, HPLC purified). SARS-CoV-2 N1 and N2 primers and probes were synthesized by IDT (oligonucleotides: 25 nmol, standard desalting; hydrolysis probe: 25 nmol, HPLC purified). Beta-actin (ACTB) primers and probe (IDT, Hs.PT.39a.22214847) were synthesized by IDT (oligonucleotides: 25 nmol, standard desalting; hydrolysis probe: 25 nmol, HPLC purified). Oligonucleotides and probes were resuspended in 10 mM Tris-HCl, pH 7.5 and 0.1 mM EDTA to 100 µM. RNase P and SARS-CoV-2 N1 primers and probes were designed for research-use only by the Centers for Disease Control and Prevention^40^. Primer and probe sequences are listed in Supplementary Table 1.

### Purification of SARS-CoV-2 patient sample

Specimens collected originally for diagnostic purposes were processed and stored in the University of California, Irvine Medical Center hospital. Under HS# 2021-8716 IRB approved protocol, samples were collected for post-diagnostic assessment from the Pathology Department at the University of California, Irvine through the Chao Family Comprehensive Cancer Center Experimental Tissue Shared Resource. Pharyngeal swabs were maintained at 4 °C until initial diagnostic test was performed, followed by freezing in the original collection tubes at -80 °C. At the time of releasing for research, the swabs were thawed and aliquoted at desired volumes, inactivated by incubation at room temperature for 30 min in 0.5% SDS^5^, and released to the investigators. A 50 µL volume of inactivated samples, in viral transport media (VTM) and 0.5% (w/v) SDS, were purified using the Zymo Research Quick-DNA/RNA viral extraction kit (D7020) following the manufacturer’s protocol and eluted in 50 µL of nuclease-free water.

### Standard RT-qPCR reactions

Standard RT-qPCR reactions were performed using either 2X qScript XLT 1-Step RT-qPCR ToughMix (Quantabio, 95132) or 4X TaqPath 1-Step Multiplex Master Mix (Invitrogen, A28522) at 1X final concentration. Forward and reverse primers were used at 500 nM with hydrolysis probes at 125 nM final concentration. Reactions were performed at either 5 µL or 10 µL after the addition of dH2O to a desired volume and 1 µL of DNA/RNA samples. Reactions were measured on a Bio-Rad CFX Connect (1855200) for 25 °C for 2 minutes, 50 °C for 15 minutes, 95 °C for 2 minutes, and 45 cycles of 95 °C for 3 seconds, and 55 °C for 30 seconds.

### RT-qPCR with SDS and Tween

RT-qPCR reactions containing SDS and Tween 80 were assembled similarly to the standard RT-qPCR reaction mixtures and contained 1X RT-qPCR master mix, 500 nM forward and reverse primers, and 125 nM of hydrolysis probes. Tween 80 was added to a desired final concentration of 3% (v/v). The SDS solution (0.1% [w/v] SDS in 10 mM Tris-HCl and 0.1 mM EDTA, pH 8.0) was added at a volume of 1 µL to the bottom of a 0.1 mL flat-top optically transparent PCR tube (Thomas Scientific, MPC-708Q) or added to the inner-side wall of the tube and allowed to dry at 70 °C for 5-10 min. Samples were added via pipetting (1 µL) or by 1 µL inoculation loop directly to the SDS solution and mixed for 15 s. Reactions were performed at 5 µL or 10 µL and volume was adjusted to consider the addition of sample by pipette or loop.

### RT-qPCR with SDS, Tween, and agarose

The SDS solution (0.1% [w/v] SDS in 10 mM Tris-HCl and 0.1 mM EDTA, pH 8.0) was added at a volume of 1 µL to the bottom of a 0.1 mL flat-top optically transparent PCR tube or added to the inner-side wall of the tube and allowed to dry at 70 °C for 5 minutes. Ultra-low gelling temperature agarose (Sigma Aldrich, A2576) was prepared at a working concentration of 2.5-5% (w/v) in dH2O by heating the mixture to 80 °C for 2 minutes and holding at 42 °C. Warm agarose was added to a RT-qPCR reaction mix (1X RT-qPCR master mix, 500 nM of forward and reverse primers, 125 nM hydrolysis probes, and 3% [v/v] Tween 80) to a final concentration of 0.5-1% (w/v). The mixture was then added to the lid or to the bottom of a 0.1 mL optically clear flat-top PCR tube and allowed to solidify at room temperature. Samples were added by pipetting (1 µL) or by 1 µL inoculation loop directly to the SDS solution and mixed for 15 seconds. Reactions were performed at 5 µL or 10 µL and volume was adjusted to consider sample addition by pipette or loop.

### RT-qPCR with SDS, Tween, and silicone oil

RT-qPCR reactions containing SDS and Tween 80 were assembled similarly to the standard RT-qPCR reaction mixtures and contained 1X RT-qPCR master mix, 500 nM forward and reverse primers, and 125 nM of hydrolysis probes. Tween 80 was added to a desired final concentration of 3% (v/v) in 5 µL. Reaction mixtures were added beneath the 5 µL silicone oil layer in a Q Tube (Quantabio, 95910-20). The SDS solution (0.1% [w/v] SDS in 10 mM Tris-HCl and 0.1 mM EDTA, pH 8.0) was added at a volume of 1 µL to the side of the tube and allowed to dry. Samples were added via 1 µL inoculation loop directly to the SDS spot and mixed for 15 seconds prior to being driven down into the silicone oil/reaction mixture at the bottom of the tube. Samples were incubated on a Quantabio Q thermal cycler (95900-4C) for 37 °C for 30 seconds, 42 °C for 4 minutes, 50 °C for 5 minutes, 95 °C for 1 minute, and 45 cycles of 95 °C for 5 seconds, and 55 °C for 10 seconds.

### RT-LAMP primer design

N1 RT-LAMP primers were designed by Huang *et al*. for the SARS-CoV-2 *N* gene^30^. Oligos were synthesized by IDT (25 nmol, standard desalting). Oligomers were resuspended in 10 mM Tris-HCl, pH 7.5 and 0.1 mM EDTA to 100 µM. RT-LAMP sequences can be found in Supplementary Table 1.

### Real-time RT-LAMP

5X RT-LAMP reaction buffer was prepared using 10X Isothermal Amplification Buffer (New England Biolabs, B0537S) with the addition of 40 mM MgSO4, 25 µM SYTO 82 (Invitrogen, S11363), 4.5% (v/v) Tween 80 and dH2O to a preferred volume. RT-LAMP reactions contained 2 µL 5X RT-LAMP reaction buffer (1X reaction buffer: 20 mM Tris-HCl, 10 mM (NH4)2SO4, 50 mM KCl, 10 mM MgSO4, 0.1% [v/v] Tween 20, 0.9% [v/v]

Tween 80, 5 µM SYTO 82, pH 8.8), 2 µL 7 mM dNTPs, 1 µL *Bst* 2.0 WarmStart DNA polymerase (8 U/µL) (New England Biolabs, M0538S), 0.25 µL WarmStart RTx reverse transcriptase (150 U/µL) (New England Biolabs, M0380S), and 1 µL 10X SARS-CoV2 N1 LAMP primer mix (2 µM F3/B3, 16 µM FIP/BIP, 4 µM LF/LB) and dH2O to 9 µL. Prior to the addition of the reaction mixture, 1 µL 0.1% (w/v) SDS in 10 mM Tris-HCl pH 8, and 0.1 mM EDTA was added to the bottom of a 0.1 mL flat-top optically transparent PCR tube. Samples were collected with 1 µL inoculation loops and mixed in the SDS solution for 15 s. After sample addition, the reaction mixture was added for a final volume of 10 µL. Reactions were monitored using a Bio-Rad CFX Connect thermal cycler at 65 °C for 50 min.

### End-point RT-LAMP fluorescent assay

End-point assays were prepared under identical conditions to real-time RT-LAMP reactions with a final concentration of 20 µM SYTO 82 for improved visualization. Reactions were incubated for 30 min and 50 min and visualized by white, green and UV (365 nm) LED lights. Images were taken by mobile phone.

## Supplementary Figures

**Supplementary Table 1:**
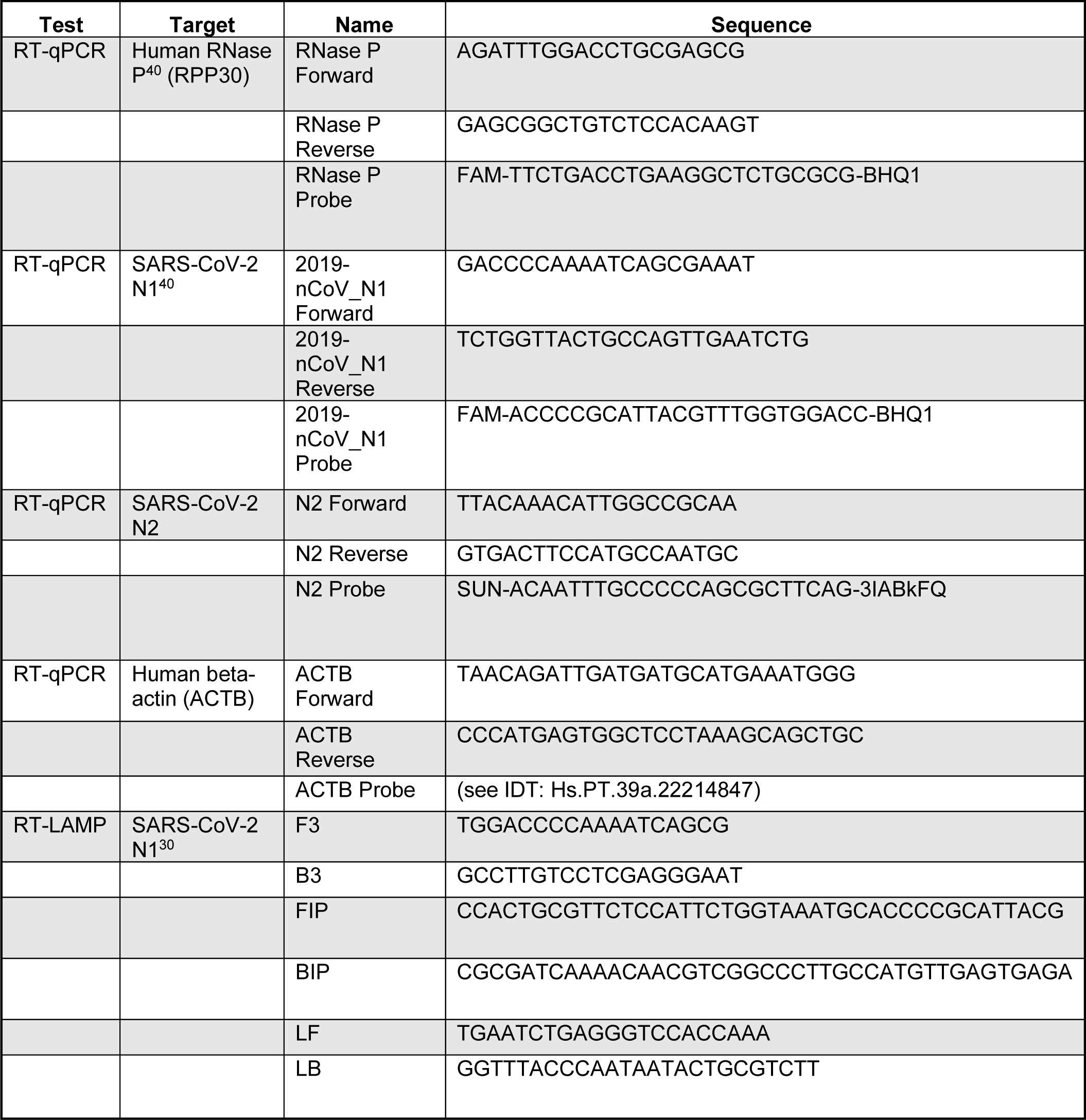
RT-qPCR and RT-LAMP primers for SARS-CoV-2. Human RNase P and SARS-CoV-2 primers and probes were designed by the Centers for Disease Control and Prevention^40^. SARS-CoV-2 N2 primers and probe were designed for this study. Human beta-actin (ACTB) primers and probe were designed by IDT (Hs.PT.39a.22214847). SARS-CoV-2 N1 RT-LAMP primers were designed by Huang *et al*^30^. All primers were purchased and synthesized by IDT.

## Notes

### Competing Interest Statement

The authors have declared no competing interest.

### Author Declarations

Under HS# 2021-8716 IRB approved protocol, samples were collected for post-diagnostic assessment from the Pathology Department at the University of California, Irvine through the Chao Family Comprehensive Cancer Center Experimental Tissue Shared Resource.

